# Human Motor Endplate Survival after Chronic Peripheral Nerve Injury

**DOI:** 10.1101/2023.10.12.23296760

**Authors:** Ranjan Gupta, Tyler R. Johnston, Vivian Y. Chen, Luigi P. Gonzales, Oswald Steward

## Abstract

**Objective:** Degeneration of motor endplates (MEPs) in denervated muscle is thought to be a key factor limiting functional regeneration after peripheral nerve injury (PNI) in humans. However, there is currently no paradigm to determine MEP status in denervated human muscle to estimate likelihood of reinnervation success. Here, we present a quantitative analysis of MEP status in biopsies of denervated muscles taken during nerve repair surgery and ensuing functional recovery.

**Methods:** This is a retrospective single-surgeon cohort study of patients (n=22) with upper extremity PNI confirmed with electromyography (EMG), treated with nerve transfers. Muscle biopsies were obtained intra-operatively from 10 patients for MEP morphometric analysis. Age at time of surgery ranged from 22-77 years and time from injury to surgery ranged from 2.5 -163 months. Shoulder range of motion (ROM) and Medical Research Council (MRC) scores were recorded pre-op and at final follow-up.

**Results:** Surviving MEPs were observed in biopsies of denervated muscles from all patients, even those greater than six months from injury. Average postoperative ROM improvement (assessed between 6-9 months post-surgery) was: forward flexion 84.3 ± 51.8°, abduction 62.5 ± 47.9°, and external rotation 25.3 ± 28.0°.

**Interpretation:** While it is believed that MEP degeneration 6 months post-injury prevents reinnervation, this data details MEP persistence beyond this timepoint along with significant functional recovery after nerve surgery. Accordingly, persistence of MEPs in denervated muscles may predict the extent of functional recovery from nerve repair surgery.

## Introduction

Peripheral nerve injuries (PNIs) often lead to significant morbidity despite medical management and surgical reconstruction. Causes of traumatic PNIs include motor vehicle crashes, stab injuries, gunshot wounds, industrial accidents, dislocations, and iatrogenic injuries.^1^ It is estimated that PNIs occur in up to 5% of all trauma patients in the United States, and 60-70% of PNIs occur in the upper extremity.^1^ PNIs are more common in males, with an average age at the time of injury ranging from 32.4-34.6 years old.^2,3^

Impairment of functional recovery post PNI is due to damage of the motor axons that innervate target muscles. Although peripheral nerve axons are capable of some regeneration, axons often must grow for long distances from the injury site to reach denervated muscles, which takes weeks or months depending on the distance.^4^ With prolonged denervation, the muscle undergoes progressive degeneration of the postsynaptic specialization containing acetylcholine receptors (the motor endplate, MEP) involving morphological regression from their normal complex, web-like pretzel form to intermediate to simple plaque structures, and eventually complete degeneration.^5^ Consequently, with extended periods of denervation, it is believed that few, if any, MEPs remain, making the denervated muscle unreceptive to reinnervation (reinnervation failure).^6^

Despite the understanding that denervated muscles undergo time dependent degeneration, physicians generally recommend a wait and see approach after PNIs with the hope that the nerve injury is a neuropraxia-type (Sunderland grade 1) and will spontaneously recover.^6^ This delay often takes the patient to the time period when nerve repair surgery is considered futile (6 months) due to expected degeneration of MEPs. For patients in which the original nerve fails to regenerate, one surgical option is a nerve transfer. Nerve transfers offer advantages as they are remote from the injury, decrease regeneration time due to MEP proximity to the donor nerve, and can lead to functional reinnervation.^7,8^ However, nerve transfers at long post-injury intervals are also considered to have a low probability of success because of the assumption that MEPs in denervated muscle have completely degenerated.

In this study, we report the results of a cohort of patients who underwent nerve transfer surgeries at different times post-PNIs with the goal of enabling reinnervation of denervated muscles. This study is based on our previous surprising discovery that MEPs persist in some patients beyond 6 months post injury, suggesting that certain patients are still candidates for surgical intervention after the 6 months post-injury period.^9^ The development of a biomarker to identify such patients could provide a means to predict success of nerve transfers/surgical interventions and overcome the current time restriction dogma to improve patient outcomes.^10^ Here, we: 1) present our method for sampling and quantitatively assessing MEP integrity from biopsies taken at the time of nerve transfers in patients with PNIs; 2) use the morphometric data to provide an initial quantitative description of MEP degeneration over time; 3) relate MEP data to postoperative recovery of range of motion (ROM) and strength.

## Materials and Methods

### Inclusion Criteria

Patients undergoing elective nerve transfer surgery for brachial plexus PNIs were enrolled under a university-approved IRB with informed patient consent to obtain muscle biopsies intra-operatively. Patients received surgery from a single surgeon between 2021-2023.

### Patient characteristics (Table 1)

Patient average age at the time of surgery was 49.6 (range 22-77 years) with a time from injury to surgery ranging from 2.5-163 months. Of the 22 patients, 2 patients received 2 concomitant nerve transfers and the other 20 received a single nerve transfer. 9 patients received an end-to-end nerve transfer while 13 received end-to-side (Table 1).

**Table 1.**
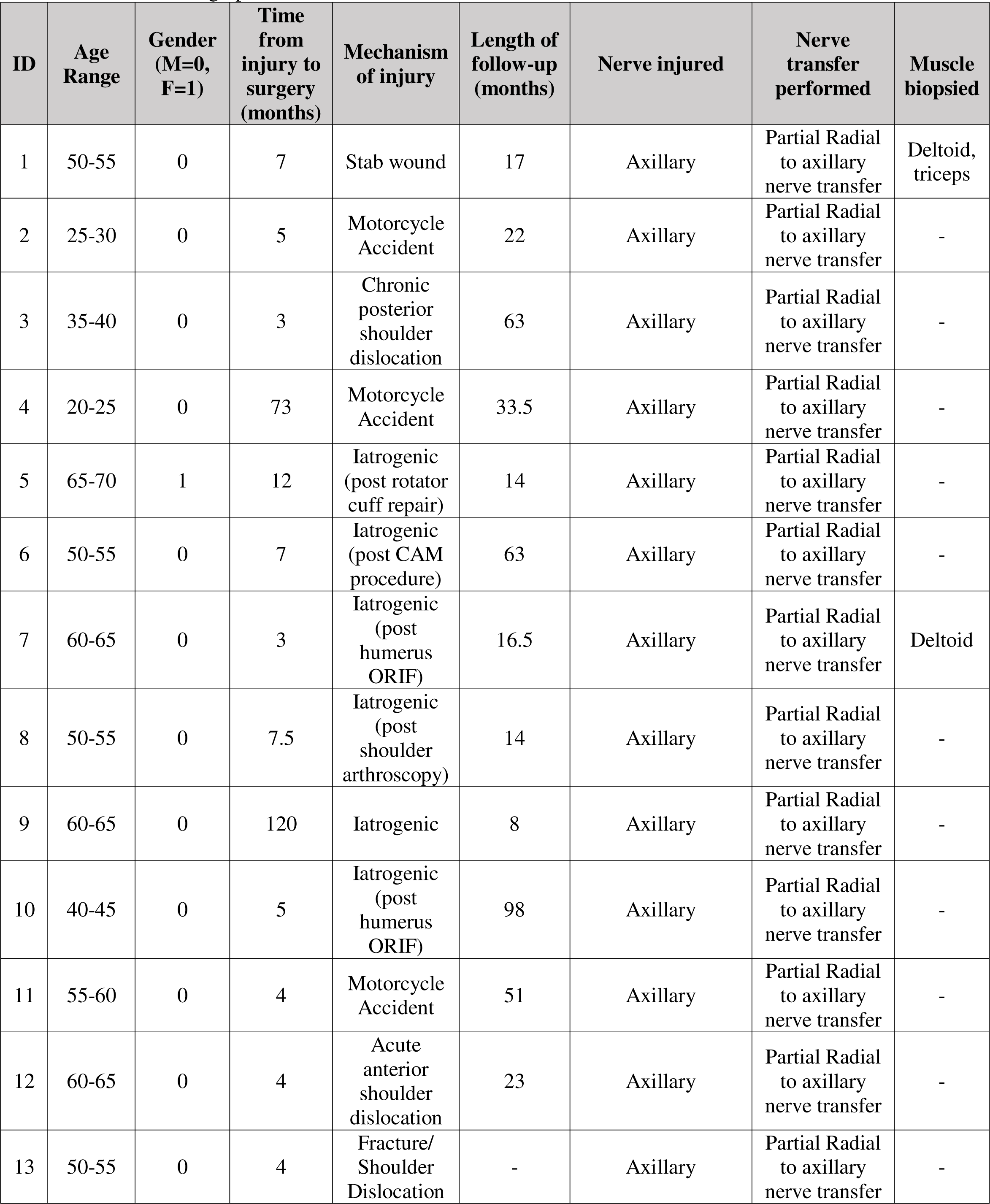
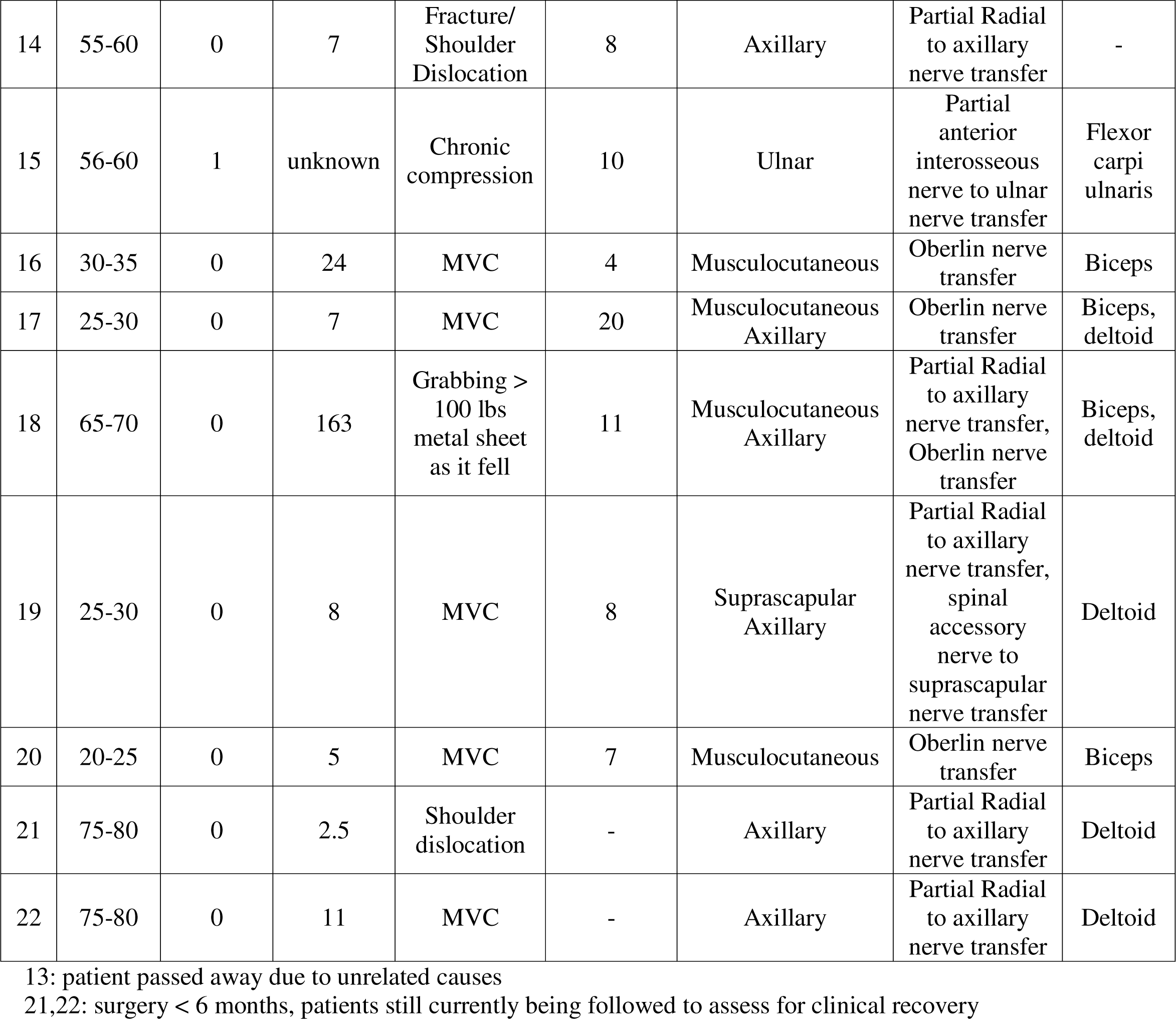
Patient demographics.

| ID | Age Range | Gender (M=0, F=1) | Time from injury to surgery (months) | Mechanism of injury | Length of follow-up (months) | Nerve injured | Nerve transfer performed | Muscle biopsied |
| --- | --- | --- | --- | --- | --- | --- | --- | --- |
| 1 | 50-55 | 0 | 7 | Stab wound | 17 | Axillary | Partial Radial to axillary nerve transfer | Deltoid, triceps |
| 2 | 25-30 | 0 | 5 | Motorcycle Accident | 22 | Axillary | Partial Radial to axillary nerve transfer | - |
| 3 | 35-40 | 0 | 3 | Chronic posterior shoulder dislocation | 63 | Axillary | Partial Radial to axillary nerve transfer | - |
| 4 | 20-25 | 0 | 73 | Motorcycle Accident | 33.5 | Axillary | Partial Radial to axillary nerve transfer | - |
| 5 | 65-70 | 1 | 12 | Iatrogenic (post rotator cuff repair) | 14 | Axillary | Partial Radial to axillary nerve transfer | - |
| 6 | 50-55 | 0 | 7 | Iatrogenic (post CAM procedure) | 63 | Axillary | Partial Radial to axillary nerve transfer | - |
| 7 | 60-65 | 0 | 3 | Iatrogenic (post humerus ORIF) | 16.5 | Axillary | Partial Radial to axillary nerve transfer | Deltoid |
| 8 | 50-55 | 0 | 7.5 | Iatrogenic (post shoulder arthroscopy) | 14 | Axillary | Partial Radial to axillary nerve transfer | - |
| 9 | 60-65 | 0 | 120 | Iatrogenic | 8 | Axillary | Partial Radial to axillary nerve transfer | - |
| 10 | 40-45 | 0 | 5 | Iatrogenic (post humerus ORIF) | 98 | Axillary | Partial Radial to axillary nerve transfer | - |
| 11 | 55-60 | 0 | 4 | Motorcycle Accident | 51 | Axillary | Partial Radial to axillary nerve transfer | - |
| 12 | 60-65 | 0 | 4 | Acute anterior shoulder dislocation | 23 | Axillary | Partial Radial to axillary nerve transfer | - |
| 13 | 50-55 | 0 | 4 | Fracture/ Shoulder Dislocation | - | Axillary | Partial Radial to axillary nerve transfer | - |
| 14 | 55-60 | 0 | 7 | Fracture/<br>Shoulder<br>Dislocation | 8 | Axillary | Partial Radial<br>to axillary<br>nerve transfer | - |
| 15 | 56-60 | 1 | unknown | Chronic<br>compression | 10 | Ulnar | Partial<br>anterior<br>interosseous<br>nerve to ulnar<br>nerve transfer | Flexor<br>carpi<br>ulnaris |
| 16 | 30-35 | 0 | 24 | MVC | 4 | Musculocutaneous | Oberlin nerve<br>transfer | Biceps |
| 17 | 25-30 | 0 | 7 | MVC | 20 | Musculocutaneous<br>Axillary | Oberlin nerve<br>transfer | Biceps,<br>deltoid |
| 18 | 65-70 | 0 | 163 | Grabbing ><br>100 lbs<br>metal sheet<br>as it fell | 11 | Musculocutaneous<br>Axillary | Partial Radial<br>to axillary<br>nerve transfer,<br>Oberlin nerve<br>transfer | Biceps,<br>deltoid |
| 19 | 25-30 | 0 | 8 | MVC | 8 | Suprascapular<br>Axillary | Partial Radial<br>to axillary<br>nerve transfer,<br>spinal<br>accessory<br>nerve to<br>suprascapular<br>nerve transfer | Deltoid |
| 20 | 20-25 | 0 | 5 | MVC | 7 | Musculocutaneous | Oberlin nerve<br>transfer | Biceps |
| 21 | 75-80 | 0 | 2.5 | Shoulder<br>dislocation | - | Axillary | Partial Radial<br>to axillary<br>nerve transfer | Deltoid |
| 22 | 75-80 | 0 | 11 | MVC | - | Axillary | Partial Radial<br>to axillary<br>nerve transfer | Deltoid |

13: patient passed away due to unrelated causes
21,22: surgery < 6 months, patients still currently being followed to assess for clinical recovery

All patients were assessed pre-operatively with electromyography (EMG) to confirm PNI. Nerve transfers performed included, but are not limited to, Oberlin, anterior interosseous nerve to ulnar nerve and radial to axillary nerve transfers (Table 1). As the sample size of patients with muscle biopsies is relatively small, an additional cohort of patients with isolated axillary nerve injuries confirmed with EMG and treated with partial radial to axillary nerve transfer by the same single surgeon between 2014-2022 was also included. This additional cohort was included to gain a more comprehensive assessment of postoperative gains in ROM and strength after nerve repair.

### Functional assessments

All patients, including those who received radial to axillary nerve transfers, but no muscle biopsies, were evaluated for muscle range of motion and Medical Research Council (MRC) scores (deltoid, triceps, wrist, and digit extension) at pre-op *and at final follow-up*. Donor morbidity was evaluated for all patients immediately postoperatively and at all follow-up visits. Statistical analyses of these functional measures were performed using the Student *t*-test.

Surgical Technique for nerve transfers: Four different types of nerve transfers within the brachial plexus were performed: Oberlin (ulnar to musculocutaneous nerve transfer), spinal accessory to suprascapular nerve transfer, AIN to ulnar nerve transfer, and a modified partial radial to axillary nerve transfer. For all procedures, long-acting paralytics and local anesthetics were not administered in order to assess for intraoperative neural function. In all cases, an intraoperative nerve stimulator was used to identify the donor fascicle. Nerve transfers were only performed when (1) there were additional redundant fascicles to preserve critical upper extremity functions, (2) appropriate donor fascicle was found within the first 60 minutes after induction of anesthesia.

For all patients, the recipient nerve was super-maximally stimulated with the intraoperative nerve stimulator to evaluate for extent of nerve injury. If there was no muscle contraction, an end-to-end transfer was performed. If there was evidence of muscle contraction (indicating that there is some nerve fiber continuity), an end-to-side transfer was performed where the recipient nerve had a perineural window created prior to anastomosis with the donor nerve. Oberlin, AIN to ulnar and spinal accessory to suprascapular nerve transfers were performed based on the technique described by Rinker et al in 2015.^11^ The modified partial radial to axillary nerve procedure was performed based on the technique described by Gupta et al in 2020.^9^

### Postoperative follow-up

Current mean follow-up in the studied cohort is 25.8 months (range, 4-98). All patients demonstrated no donor site defect in the immediate post-operative unit. All patients received repeated donor nerve strength testing at subsequent follow-up visits. There were no cases of donor site morbidity. No patient required subsequent revision surgery.

#### Muscle biopsy harvest

Muscle biopsies were harvested from denervated muscles intraoperatively. In 7 patients, biopsies were from the deltoid muscle (Table I); in 4 cases, biopsies were from the biceps and in one patient, the biopsy was from the flexor carpi ulnaris. Muscle biopsies were taken from the periphery of the muscle, immediately adjacent to where surgical retractors were placed. Due to retractor placement, these muscle samples were would have otherwise been discarded. Thus, this location of harvest ensured minimal patient morbidity. Approximately 2-3 2×2×2 cm muscle biopsies were taken from each patient, depending on degree of muscle atrophy. In patients with less muscle bulk, only 2 samples were obtained. In patients with more muscle bulk, 3 or more samples were taken. After biopsy, each muscle sample was placed in a sterile container and transported on ice directly from the operating room to our laboratory for processing. All biopsies were processed the same day as harvest.

### Tissue fixation and immunostaining

Muscle biopsies from all patients were evaluated for MEP morphometric analysis. Muscle samples were first evaluated using a dissecting microscope (magnification 10x) to separate fatty tissue from the muscle fibers. The muscle was then divided into 1cm portions. Muscle tissue was processed without sectioning, thereby preserving muscle fiber architecture (figure 1). Tissue clearing reagents and protocol were based on procedures described by Muniterfering et al, 2018.^12^ Muscle biopsies were fixed in 4% para-formaldehyde (PFA) overnight. After washing with phosphate buffered saline (PBS), biopsies were placed in clear, unobstructed brain/body imaging cocktails and computational analysis (CUBIC) R1 solution (urea, quadrol, Triton-x-100). Tissue samples were incubated at 37°C on a rotating mixer until the tissue turned clear or light yellow in color. Each sample took 10-14 days before turning light yellow after CUBIC R1 immersion, changing CUBIC solution every 2 days. Human samples rarely turned completely clear even following prolonged CUBIC R1 incubation. After clearing, samples were washed 3X (two hours each) in CUBIC IHC buffer, followed by an overnight wash in CUBIC IHC buffer (bovine serum albumin, triton-x, sodium azide). Primary antibodies were then added. All samples were stained with neurofilament (Biolegend Cat #837904 RRID: AB_2566782), synaptophysin and either AChR α1 (Novus Biologicals Cat#NBP2-81070), β (AbcamCat# ab76159) or δ (Novus Biologicals Cat#NB120-2804). After incubation in primary antibodies, samples were washed 3X (two hours each) in CUBIC IHC buffer, followed by an overnight wash in CUBIC IHC buffer. Samples were then incubated in secondary antibodies in CUBIC IHC buffer, either: (1) Donkey anti-Mouse IgG (H+L) Highly Cross-Adsorbed Secondary Antibody, Alexa Fluor Plus 488 (Thermofisher Cat A32766, RRID: AB_2762823), (2) Goat anti-Rabbit IgG (H+L) Highly Cross-Ad-sorbed Secondary Antibody, Alexa Fluor Plus 594 (Thermofisher Cat# A32740, RRID: AB_2762824) or (3) Goat anti-Rabbit IgG (H+L) Highly Cross-Adsorbed Secondary Anti-body, Alexa Fluor Plus 488 (Thermofisher Cat# 32731, RRID:AB_2633280). The samples were washed 3X (two hours each) plus an additional overnight wash in CUBIC IHC buffer and then mounted for imaging.

**Figure 1.**
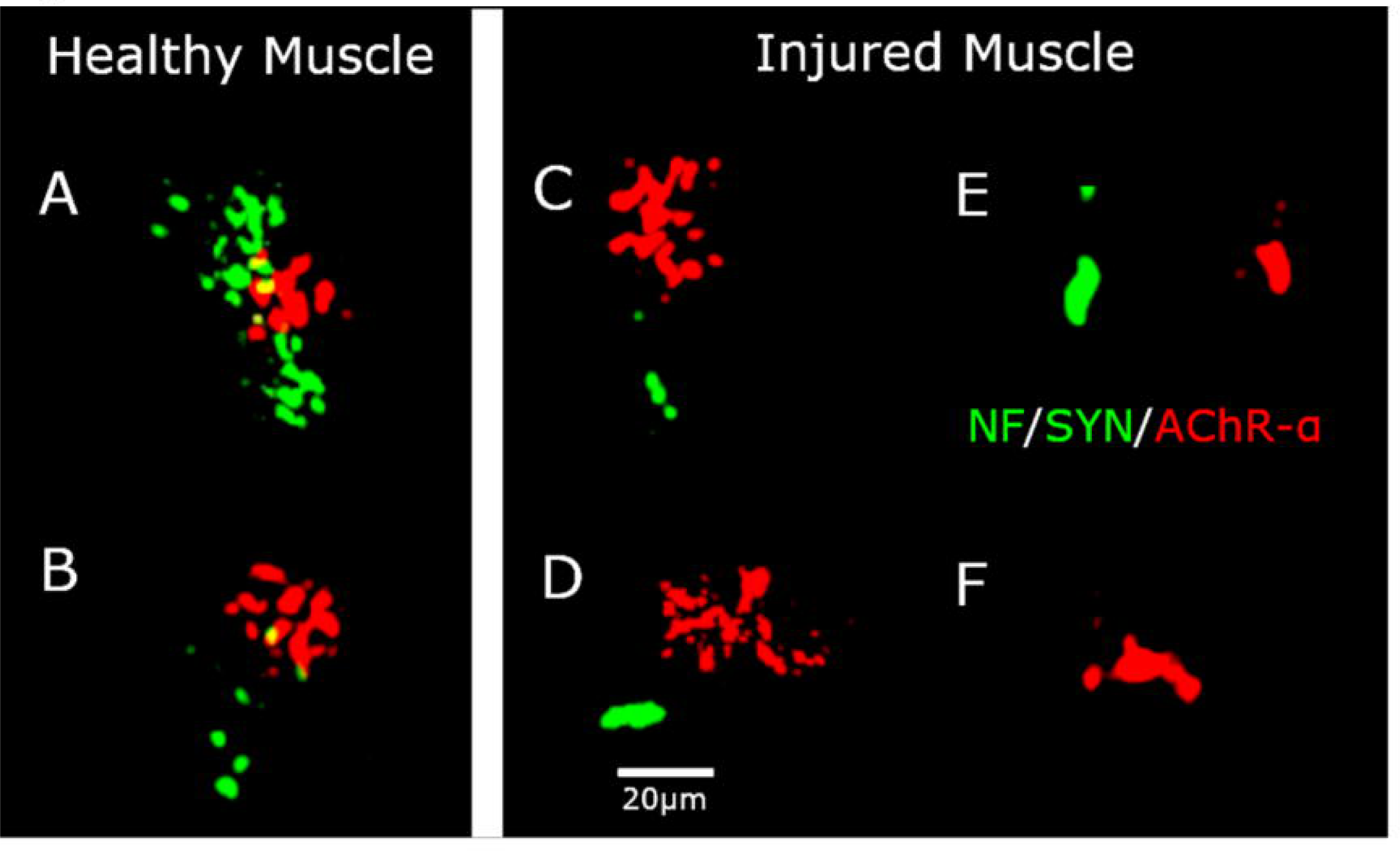
MEP morphology. lmmunofluorescence of MEPs at 20x magnification. Red signal represents MEPs and is stained with AChR-a. Green signal represents the nerve terminal and is stained with neurofilament/synaptophysin.(A-B) Healthy pretzel MEPs as seen in uninjured muscle. Healthy MEPs contain a compact structure with multiple perforations. (C-F) unhealthy MEPs in injured muscle. (C-D) intermediate MEPs are larger and more spread out with relatively complex structures. (E-F) plaque MEPs are small and simple when compared to intermediate and pretzel MEPs.

### Tissue imaging

After mounting, muscle samples were imaged with a Keyence BZ-X810 inverted fluorescence phase contrast microscope. Biopsies were first scanned at 4x magnification to determine MEP distribution and number. Each individual MEP was then examined at 10x and then all MEPs were imaged at 20x magnification. MEP morphology was quantified using BZ-X10 analyzer and ImageJ assessing number of perforations, MEP area, and overall shape. MEP form was classified by two independent investigators as “normal pretzel, intermediate, or simple plaque” according to criteria defined by Chao et al^5^

## Results

In our muscle biopsy cohort, 10 patients met inclusion criteria—1 female and 9 males. An additional 12 patients were treated with radial to axillary nerve transfers following isolated axillary nerve injuries (but had no muscle biopsies); these are included in ROM and MRC analysis—11 male and 1 female (Table 1) (22 patients total).

### MEP analysis

MEPs were present in biopsies from all 10 patients, even the 6 patients with greater than 6 months from injury to biopsy. However, the MEPs that were present exhibited clear signs of degeneration in all patients (Figure 2). Average percentage of abnormal MEPs (plaque and intermediate morphologies) was 87.3% (range, 61.5-100%). Average percentage of healthy MEPs (pretzel morphology) was 10.3% (0-20%). Average number of MEPs in each biopsy sample was 16.3 (range, 5-37). Average percentage of MEPs with intact nerve terminals was 45.4% (range, 0-100) (Table 2). One patient (#20) had below average percentages for both categories. Two other patients (#1 and #19) had below average healthy MEPs, but average/above average innervated MEPs.

**Figure 2.**
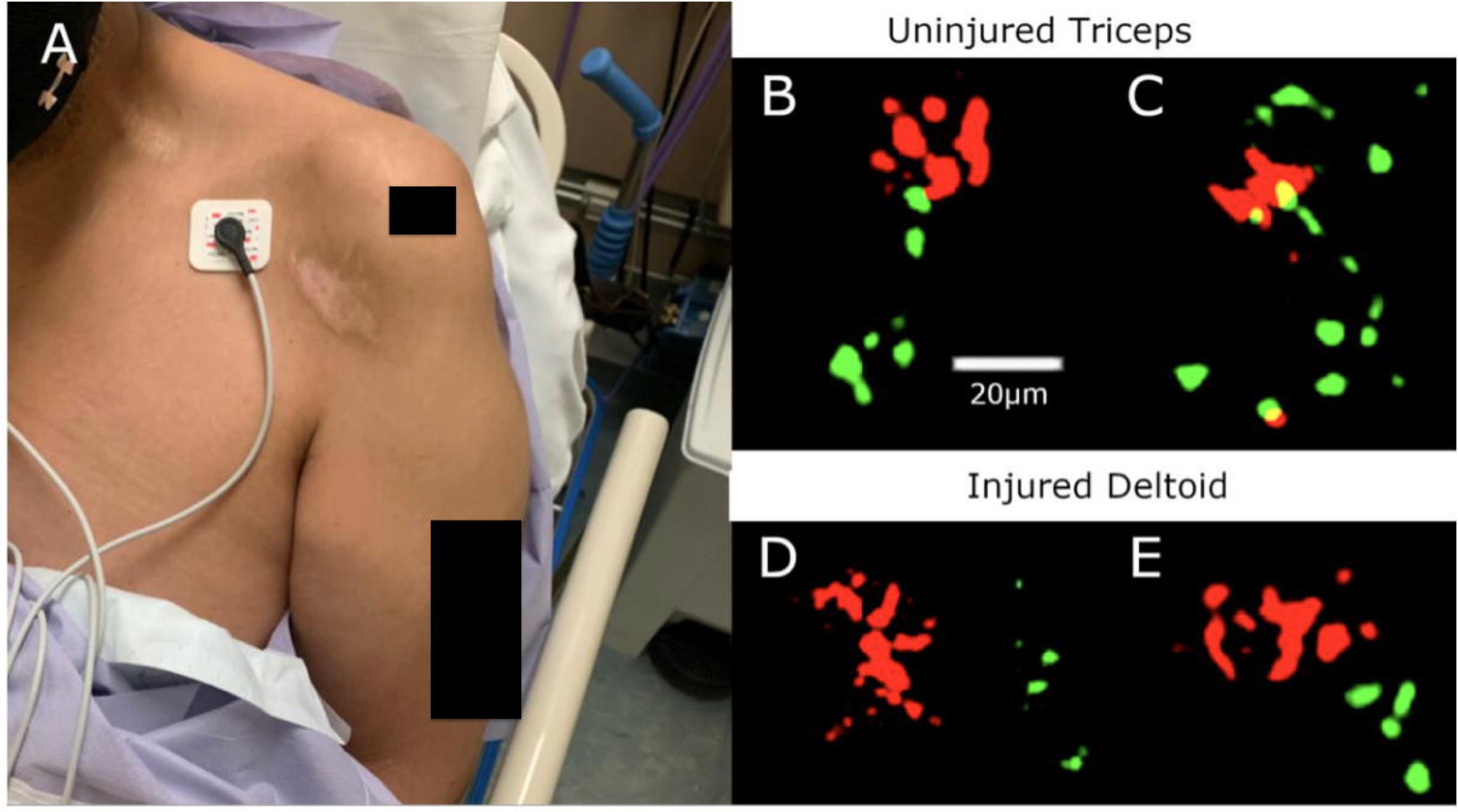
Muscle data from patient with isolated axillary nerve injury. (A) Gross atrophy of deltoid is apparent on clinical exam at 7 months post-injury. (B-E) Immunofluorescence of MEPs at 20x magnification. Red signal represents MEPs and is stained with AChR-a. Green signal represents the nerve terminal and is stained with neurofilament/synaptophysin. (B-C) Pretzel MEPs in uninjured triceps with intact nerve terminal. (D-E) Intermediate MEPs in injured deltoid.

**Table 2.** Analysis of MEPs from human muscle biopsies.

| **ID** | **Gender** | **Age Range** | **Isolated Nerve Injury (1) or Multiple Nerve Injuries (2)** | **Nerve(s) injured** | **Time from Injury to Surgery/ Biopsy (months)** | **Percentage of healthy MEPs (pretzel)** | **Percentage of unhealthy MEPs (intermediate + plaque)** | **Percentage of Innervated MEPs** | **Total number of MEPs** |
| --- | --- | --- | --- | --- | --- | --- | --- | --- | --- |
| 1 | M | 50-55 | 1 | Axillary | 7 | 0 | 100 | 100 | 5 |
| 7 | M | 60-65 | 1 | Axillary | 4 | 20 | 80 | 0 | 5 |
| 15 | F | 55-60 | 1 | Ulnar | unknown | 20 | 80 | 60 | 5 |
| 16 | M | 30-35 | 2 | Musculocutaneous | 24 | 4.2 | 95.8 | 37.5 | 24 |
| 17 | M | 25-30 | 2 | Musculocutaneous<br>Axillary | 7 | 15.4 | 84.6 | 69.2 | 13 |
| 18 | M | 65-70 | 2 | Musculocutaneous<br>Axillary | 163 | 16.7 | 83.3 | 58.3 | 12 |
| 19 | M | 25-30 | 2 | Suprascapular<br>Axillary | 8 | 5.4 | 94.6 | 46.0 | 37 |
| 20 | M | 20-25 | 1 | Musculocutaneous | 5 | 6.1 | 93.9 | 39.8 | 33 |
| 21 | M | 75-80 | 1 | Axillary | 2.5 | 15.4 | 61.5 | 30.8 | 13 |
| 22 | M | 75-80 | 2 | Axillary | 11 | 0 | 100 | 12.5 | 16 |

There was a weak but statistically significant negative correlation between age and number of MEPs (r: -0.35, p=0.05) (figure 3). Age was otherwise not correlated with any other factors.

**Figure 3.**
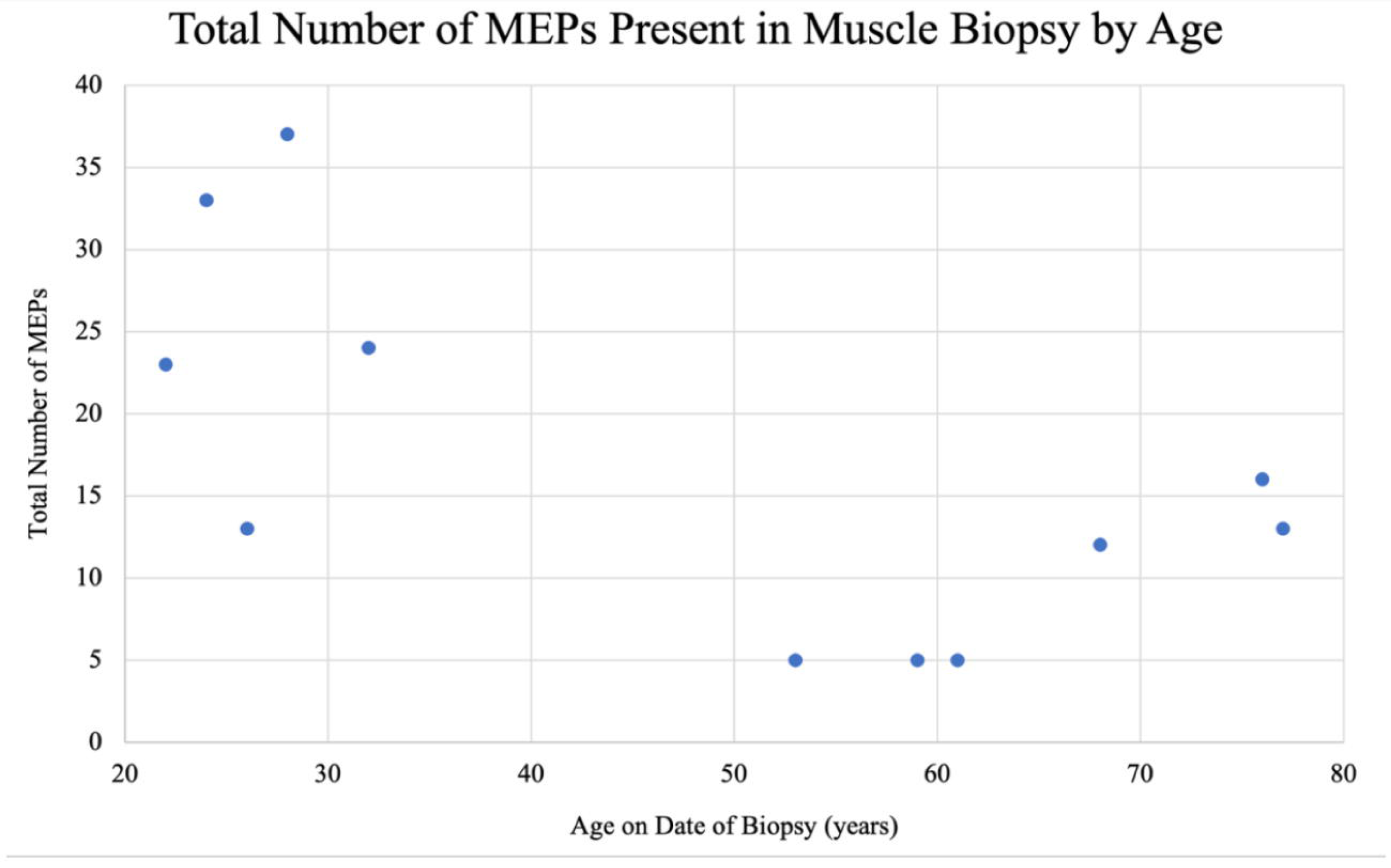
Scatterplot depicting correlation between age (on surgery date) and the total number of motor end plates (MEPs). Age was inversely correlated with number ofMEPs (r: -0.35, p=0.05).

There was no correlation between time from injury to surgery and percentage of healthy MEPs (r: 0.05, p=0.42), time from injury to surgery and percentage of abnormal MEPs (r: -0.02, p=0.82), time from injury to surgery and percentage of innervated MEPs (r: 0.10, p=0.64) and time from injury to surgery and number of MEPs (r: -0.03, p=0.70). After performing an ANOVA analysis, there was no statistically significant difference between isolated or multiple nerve injuries in the percentage of pretzel MEPs, percentage of abnormal (intermediate and plaque) MEPs, percentage of innervated MEPs, and number of MEPs (p=0.82).

There was no correlation between percentage of pretzel MEPs and postoperative gain in ROM (r: 0.66, p=0.86), percentage of abnormal (intermediate and plaque) MEPs and postoperative gain in ROM (r: -0.66, p=0.82) or percentage of innervated MEPs and postoperative gain in ROM (r: 1.00, p=0.19).

Based on his clinical course, one patient received three surgeries and 3 biopsies (Figure 4). The pre-nerve transfer biopsy contained no pretzel MEPs whereas biopsies at 7- and 15-months post-transfer revealed 62.5% and 66% of MEPs with pretzel morphology, respectively (Table 2).

**Figure 4.**
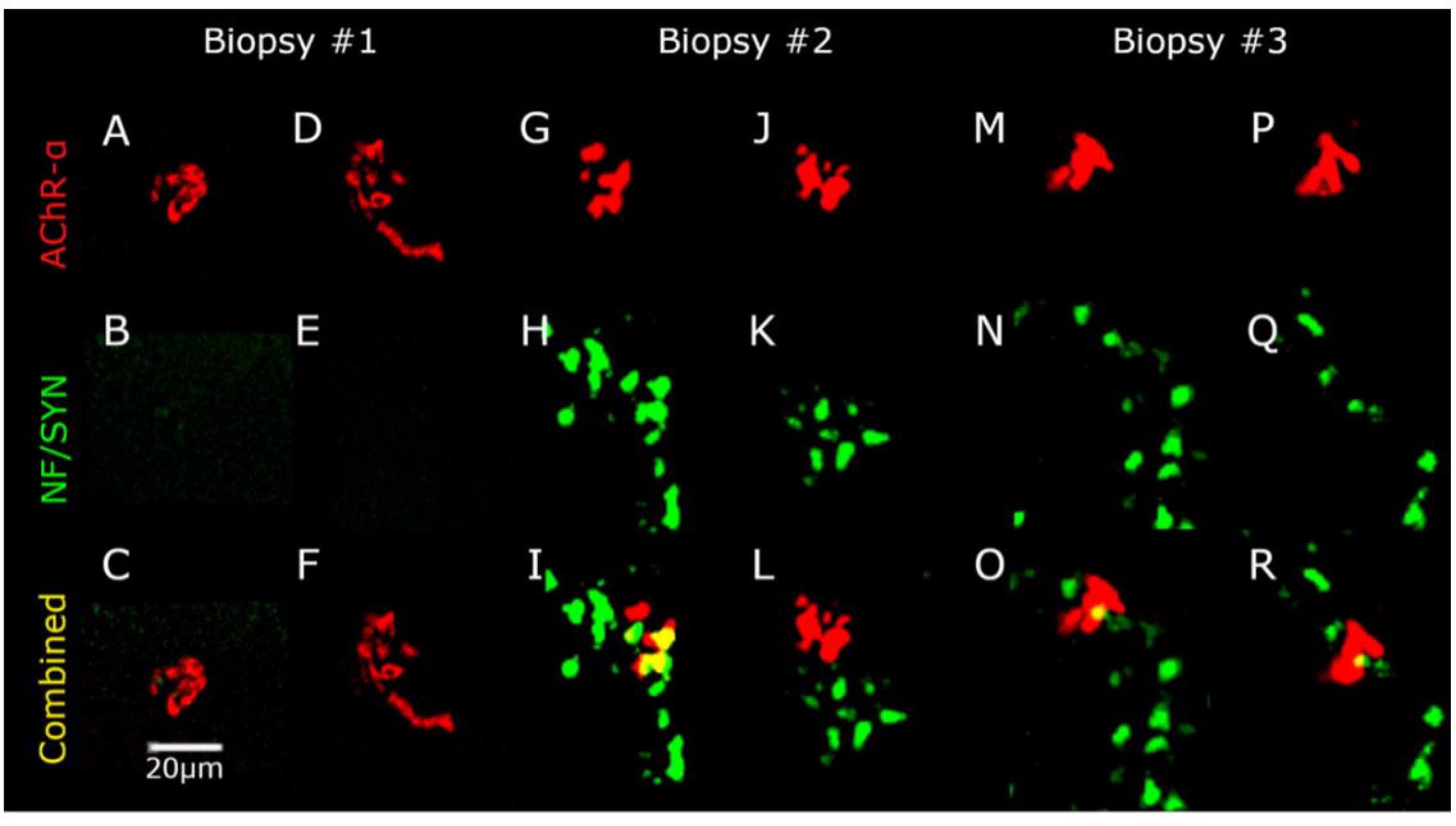
MEPs from patient with isolated axillary nerve injury who received three sequential biopsies. Red signal represents MEPs and is stained with AChR-a. Green signal represents the nerve terminal and is stained with neurofilament/synaptophysin. (A-F) MEPs from pre-nerve transfer biopsy demonstrating absence of nerve terminals. (G-L) MEPs from 7 months post­ nerve transfer biopsy demonstrating rescue of nerve terminals and MEP morphology. (M-R) MEPs from 15 months post-nerve transfer biopsy demonstrating maintenance of healthy MEPs with intact nerve terminals.

Furthermore, the percentage of MEPs with an intact nerve terminal improved from 0% pre-transfer to 75% and 83% at 7- and 15-months post-nerve transfer, respectively.

### Electromyography

Of the 22 patients, 19 had accessible detailed EMG reports confirming PNI. The remaining 3 patients only had brief notes with abbreviated EMG results, 1 from the muscle biopsy cohort, the other 2 from the nerve transfer only cohort. One cardinal sign of denervation is the appearance of fibrillation potentials, which typically appear in denervated muscles 1-4 weeks post-nerve injury^13^; 1+ fibrillation potentials indicate one or two potentials in at least two areas, while 4+ fibrillations indicate diffuse persistent discharges across.^14^ Increasing fibrillation potential grades correlate with increased severity of nerve damage.^15^ Of the muscle biopsy cohort, 1 patient had 4+ fibrillation potentials, 7 had 3+ fibrillation potentials, and 1 had 1+ fibrillation potentials. Distribution of fibrillation potentials for the other cohort of patients was: 2 had 3+ potentials, 7 had 2+ potentials and 1 patient had 1+ potentials.

EMG studies also provided data regarding other electromyographical abnormalities indicating denervation or signs of reinnervation. Motor unit recruitment is often used to map out severity of nerve lesions— the greater the reduction in recruitment, the more severe the nerve injury.^16^ 9/10 patients in the muscle biopsy cohort had significant reductions in motor unit recruitment, and 1 had zero motor unit recruitment. A similar distribution was also present in the nerve transfer only cohort: 10 patients had reduced recruitment, 1 had single unit recruitment and 1 had zero motor unit recruitment.

Another measure involves motor unit potentials which assess innervation status. Zero motor unit potential represents severe muscle atrophy and neural degeneration.^17^ Reinnervation is evidenced by long duration, high amplitude motor unit potentials. In the muscle biopsy cohort, 8 had long duration, high amplitude motor unit potentials and 2 had zero motor unit potential. In the nerve transfer only cohort, 6 patients had long duration, high amplitude motor unit potentials, 2 had normal duration motor unit potentials and 2 had zero motor unit potentials.

In addition to motor unit potentials, nascent potentials are markers of reinnervation. Nascent potentials are low amplitude, polyphasic signals that appear only when axonal regeneration has begun. Thus, nascent potentials are only present in patients several months post-injury.^18^ Initial nascent potentials are small and incapable of generating clinically detectable movements but represent early signs of reinnervation.^18^ 0 patients in the muscle biopsy cohort had nascent potentials, while 6 patients in the nerve transfer only cohort had detectable nascent potentials, suggesting some early stage reinnervation in these patients.

2 patients in muscle biopsy cohort had postoperative EMGs. One patient (#20) was 4 months post-op and the other (#7) was 5.5 months post-op. Both patients exhibited signs of increased innervation by EMG, evidenced by increased polyphasic units, appearance of nascent potentials and decreased fibrillation potentials.

### ROM and Strength testing (Figure 5)

Mean preoperative MRC score was 2. This improved to a postoperative mean MRC score of 4+. Average postoperative ROM improvement (assessed between 6-9 months post nerve transfer) *was*: forward flexion 84.3 ± 51.8°, abduction 62.5 ± 47.9°, and external rotation 25.3 ± 28.0°. Active ROM data was stratified by transfer type: end-to-end (Group A) and end-to-side (Group B), see Table 3. From pre-op to final follow-up, Group A mean gain in forward flexion was 65.6 ± 43.4°; Group B was 109.1 ± 55.1° (p=0.12). Group A mean gain in abduction was 48.1 ± 36.5°; Group B was 81.2 ± 57.6° (p=0.21). Group A mean gain in external rotation was 18.1 ± 29.4°; Group B was 35 ± 25.2 (p=0.28)°.

**Figure 5.**
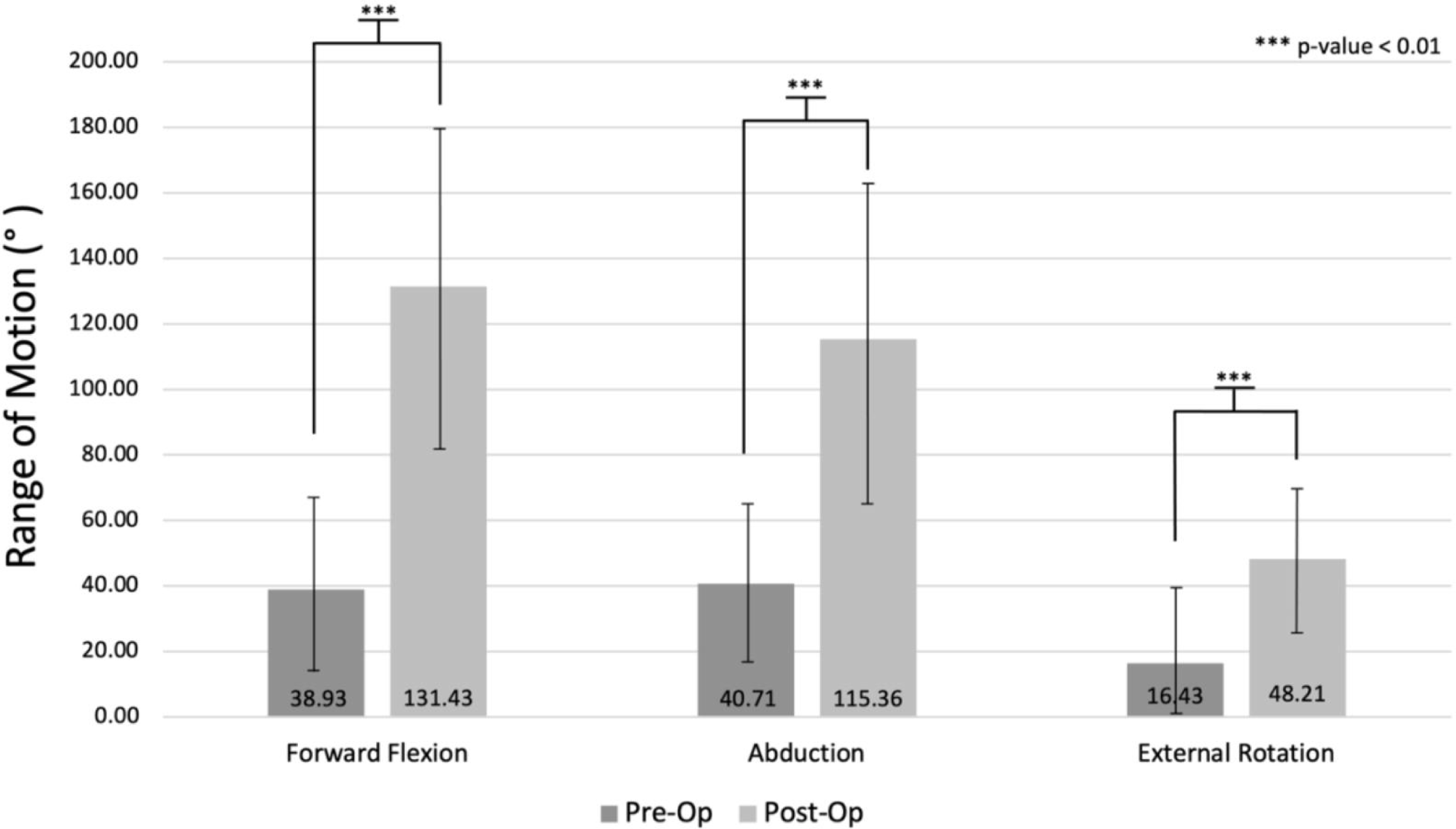
Mean pre- and post-operative range of motion.

**Table 3.** Range of motion of patients before and after nerve transfer.

| Study ID | Gender (M=0; F=1) | End-to-End (A) or End-to-Side (B) | Pre Nerve Transfer ROM (°) |  |  | Post Nerve Transfer ROM (°) |  |  |
| --- | --- | --- | --- | --- | --- | --- | --- | --- |
|  |  |  | Forward Flexion | Abduction | External Rotation | Forward Flexion | Abduction | External Rotation |
| 1 | 0 | A | 35 | 35 | 0 | 120 | 110 | 75 |
| 2 | 0 | A | 30 | 15 | 10 | 125 | 85 | 60 |
| 3 | 0 | A | 35 | 35 | 0 | 160 | 140 | 60 |
| 4 | 0 | A | 35 | 35 | 0 | 170 | 150 | 45 |
| 5 | 1 | A | 20 | 80 | 30 | 90 | 90 | 35 |
| 6 | 0 | A | 110 | 90 | 60 | 110 | 90 | 60 |
| 8 | 0 | B | 35 | 35 | 0 | 180 | 180 | 60 |
| 9 | 0 | B | 35 | 35 | 0 | 180 | 160 | 60 |
| 11 | 0 | B | 45 | 45 | 60 | 180 | 170 | 70 |
| 12 | 0 | B | 50 | 50 | 10 | 160 | 150 | 45 |
| 14 | 0 | B | 25 | 25 | 0 | 135 | 110 | 45 |
| 18 | 0 | A | 5 | 5 | 0 | 85 | 45 | 0 |
| 19 | 0 | A | 80 | 80 | 60 | 140 | 130 | 60 |
| 20 | 0 | B | 5 | 5 | 0 | 5 | 5 | 0 |

Patients excluded from ROM analysis:
7, 10: proximal humerus nonunion
13: patients passed away due to causes unrelated to surgery
15: no shoulder ROM analyzed, only grip strength
16: severe traumatic brain injury
17: no shoulder ROM analyzed, only biceps strength
21,22: insufficient follow up due to recent surgery

One patient (#15) received AIN to ulnar nerve transfer, thus the shoulder ROM and MRC was not evaluated. The patient’s grip strength did increase from 17 pounds pre-op to 35 pounds post-op. One patient (#17) only received an Oberlin nerve transfer, him shoulder ROM and MRC was also not evaluated. His biceps strength to increase from M1 to M3-4 postop.

6 patients were excluded from ROM and MRC score analysis due to other confounding variables: 1 patient passed away from causes unrelated to the procedure, 1 patient sustained a severe traumatic brain injury, 2 patients had insufficient follow up time period (< 6 months post op) to assess for functional recovery, 2 patients had non-unions of proximal humerus fractures which significantly limited their shoulder motion and strength. However, following restoration of deltoid function with nerve transfer, one of these fracture non-unions was successfully treated with reverse total shoulder arthroplasty.

### MEPs, EMGs and postoperative recovery

Two patients in the muscle biopsy cohort had both pre and post nerve transfer EMGs. Both patients had MEP persistence at the time of the initial muscle biopsy. One patient (#20) had received his EMG four months post op due to lack of clinically significant recovery. This is the aforementioned patient with low numbers of healthy MEPs (6%) and innervated MEPs (39.4%). This patient also had evidence of severe high-grade injury on preoperative EMG with zero motor unit recruitment and zero motor unit potential. This patient’s post-operative EMG demonstrated interval improvement (appearance of nascent potential and decreased fibrillation potentials), but there was no clinically meaningful functional improvement.

Another patient (#7) received 2 EMGs and 3 sequential biopsies. This patient’s initial biopsy demonstrated above average percentage of healthy MEPs (20% vs avg 10.3%), but a below average percentage of innervated MEPs (0% vs 45.5%). This patient’s second EMG (5.5 months post op) also demonstrated interval improvement. Furthermore, each subsequent biopsy demonstrated improvements in MEP morphology and innervation status after undergoing nerve transfer.

## Discussion

Given findings of early MEP degeneration in amyotrophic lateral sclerosis (ALS) and spinal muscular atrophy (SMA) that proceeds in a retrograde manner with degeneration of the distal motor axon and eventual loss of neuronal cell bodies, understanding human MEP degeneration will accelerate discovery not only in acute traumatic nerve injury but also across a wide range of progressive neurodegenerative diseases as we seek to better understand the unique molecular interplay of human nerve/muscle interactions.^19–22^ This study extends our previous report that MEPs can persist in denervated human muscles at long intervals after PNI and that patients with surviving MEPs exhibit meaningful functional recovery after delayed nerve transfer reconstructive surgery.^9^ This finding further motivates development of objective, quantitative assessments of functional recovery potential after surgical nerve repair.

In our cohort of 10 patients with muscle biopsies, MEPs persisted in all patients, although morphometric analysis revealed significant deterioration of normal MEP structure. Nine of the 10 patients also exhibited significant post-surgery improvements in function, even the two patients who did not undergo nerve transfer until > 120 months post-injury. This finding further supports our working hypothesis that the presence of MEPs in denervated muscle at long post-injury intervals, even MEPs with abnormal structure, suggests that patients may be favorable candidates for nerve transfer, despite being beyond the accepted treatment window. There was no correlation between extent of MEP degeneration and time from injury to biopsy, suggesting that rather than there being a standard timeline for loss of reinnervation competence, MEP preservation may be patient-specific. If so, objective criteria to assess reinnervation competence are of potential major significance for both surgeons and patients considering this elective surgical procedure at long post-injury intervals.

In the present cohort of 10 patients, MEP degeneration status was not significantly related to the extent of postoperative recovery. Of note, however, younger age was associated with a higher number of MEPs. This suggests that MEP preservation can be a potential factor explaining why younger patients tend to have better postoperative recoveries when compared to older patients. Perhaps because more MEPs are preserved in the denervated muscle, serving as a ‘baby-sitter’ for the post-synaptic acetylcholine receptors to retain their architecture, a greater number of functional connections can subsequently be established by reinnervating axons after reconstructive surgery, resulting in better functional outcomes. However, additional studies will be required to assess this hypothesis.

In addition to MEP numbers, the status of the MEP, specifically the morphology and innervation status was not significantly related postoperative recovery; however, this may be due to our relatively small sample size. Our one individual with the most significant signs of MEP degeneration (#20) was the only patient of the muscle biopsy cohort to not experience clinically significant recovery. While his 4 months postoperative EMG demonstrated early physiological evidence of nerve reinnervation, no clinically significant recovery was apparent, indicating that his MEPs were still receptive towards reinnervation; however, perhaps not enough functional MEPs had been preserved distally for clinically significant recovery. Future studies will be required to expand upon how the parameters of MEP morphometrics correlate with clinical outcomes/functional recovery potential.

Of note, the patients with multiple surgeries and associated biopsies provided evidence that improvement in ROM was associated with recovery of MEP measures. In patient (#7) with serial biopsies, the second and third biopsies demonstrated improvement of MEP characteristics when compared to the first biopsy. This suggests that reinnervation enabled by surgical repair rescued MEP morphology. These data from a single patient suggest that the status of the target muscle is related to clinical recovery.

The current literature on postoperative outcomes of nerve transfers for the upper extremity exhibits significant variability. Desai et al.^23^ retrospectively evaluated 27 patients who received the nerve transfer for either brachial plexus injury or isolated axillary nerve injury. Out of 27 patients, 4 achieved M5 score, while 13 achieved M4 score, and average postoperative abduction following the procedure was 114°. In Wells et al.’s^24^ systematic review, 18 out of 30 patients with axillary nerve injuries had clinical improvement postoperatively following radial nerve transfer (defined by the researchers as an increase greater than 40° in shoulder abduction and M3 or above strength). However, Wells et al.’s definition of clinical improvement may not translate to postoperative functional recovery as patients with M3 score and < 100° abduction will not be able to perform any overhead activities. Thus, while most studies document some clinical improvement, the results remain variable. As neither study included preoperative EMG or MEP status, the variability in postoperative outcomes remains a mystery in these studies. In contrast, our study was able to assess the status of the target end-organ. In doing so, we were able to better predict patient outcomes. While we still had variable postoperative results, the variability can be correlated with the target-end organ muscle status--those with high grade degeneration were more likely to experience poor functional outcomes. Accordingly, our study suggests that because there are differences in timeline of MEP degeneration between each patient, assessing both EMG and MEP degeneration can better predict the individual patient functional recovery.

In summary, all patients in our study displayed gross preoperative atrophy of denervated muscles with significantly abnormal EMGs and clinical findings consistent with peripheral nerve injury (without meaningful reinnervation or clinical recovery). Of the 15 patients included in our ROM/MRC analysis, 13 (86.7%) exhibited significant improvements post-op. While the other 2 patients (#6, #20) did not recover additional ROM or strength, neither patient experienced donor nerve site morbidity or worsened ROM/strength at the shoulder. The poor functional recovery of these 2 patients is unsurprising as both patients had evidence of significant, high-grade peripheral nerve injuries with severe degeneration of the target end organ based upon their respective preoperative EMGs and muscle biopsy results. Thus, in our cohort of patients, objective preoperative assessments were able to successfully predict the poor postoperative outcomes, emphasizing the importance of these objective preoperative markers. The overall impressive recovery of ROM and MRC scores following nerve transfer suggests that, regardless of time from injury to surgery, patients across a broad range of ages and nerve injuries are candidates for reinnervation surgery, provided their target end organ does not demonstrate severe degeneration.

The lack of an objective method to assess and predict outcomes post nerve repair makes it difficult to ascertain which patients are favorable surgical candidates. While we currently have surrogate markers, such as physical exam and EMGs, no consistent objective marker exists to assess the target end organ directly. Accordingly, our methodology can fill this void and provide valuable preoperative insight. By obtaining a pre-operative biopsy that can quantitatively and qualitatively assess the MEPs present in the sample, the possibility of successful reinnervation can be better ascertained.

The strengths of this study include the variety of injury characteristics, broad patient demographics, consistent surgical technique, and objective measurements of target end-organ response. Our study is one of few to measure the structural response of MEPs after nerve reinnervation. Our study is also one of the few studies to incorporate clinical, electrophysiological and objective data into our analysis. Additionally, all the surgeries included were performed by a single surgeon, eliminating surgeon-specific factors that may confound results.

Limitations to our study include its retrospective nature, small sample size, lack of a control group, and variation in ROM data. While our study is retrospective, all patients had rigorous follow-up with well documented pre- and post-operative data. Our muscle biopsy is also a current ongoing, prospective study; however, all patient follow up data is collected and analyzed retrospectively. Our sample size is limited due to the relatively low incidence of peripheral nerve injuries;^1^ however, we collected data over a period of eight years to accumulate a larger cohort of patients. Furthermore, our mean pre-operative versus final follow-up ROM demonstrated a large effect size, increasing the statistical power of our study.

PNIs are difficult to manage due to numerous factors that make it difficult to determine functional recovery potential. Patients can often be deemed as unsuitable candidates for surgical repair due to delayed time from injury to clinic/hospital presentation, thus missing “the critical window.” However, our patients’ drastic improvements in ROM, strength, and persistence of target end-organ MEPs suggest that the preconceived belief of a universal timepoint from injury for which MEPs degenerate is incorrect. Rather, the timeline of MEP degeneration is individualized, necessitating an objective preoperative marker to better evaluate each patient’s recovery potential. In conclusion, the results of our study suggest that (1) functional recovery post PNI is possible regardless of the timing of surgery and (2) MEP morphometric analysis from preoperative muscle biopsies is a promising preoperative test to evaluate postoperative recovery potential.

## Acknowledgements

This work was supported by NIH NIA AG081739 (RG).

## Author Contributions

- Conception and Design of the Study: RG, TJ, OS
- Acquisition and Analysis of Data: RG, TJ, VC, LG
- Drafting of Manuscript and Figures: RG, TJ, OS, VC, LG

### Data Availability

Anonymized data not published within this article will be made available by request from any qualified investigator.

